# Safety, Virology, Pharmacokinetics, and Clinical Experience of High-dose Intravenous Sotrovimab for the Treatment of Mild to Moderate COVID-19: An Open-label Clinical Trial

**DOI:** 10.1101/2023.02.02.23285352

**Authors:** Jaynier Moya, Marisol Temech, Sergio Parra, Erick Juarez, Reinaldo Hernandez-Loy, Juan C. Moises Gutierrez, Jorge Diaz, Rubaba Hussain, Scott Segal, Claire Xu, Andrew Skingsley, Gretja Schnell, Asma El-Zailik, Jennifer E. Sager, Melissa Aldinger, Elizabeth L. Alexander, Gerard Acloque

## Abstract

**Background:** 500 mg intravenous (IV) sotrovimab has been shown to be well tolerated and efficacious against pre-Omicron strains in treating patients with mild to moderate coronavirus disease 2019 (COVID-19) at high risk for disease progression.

**Methods:** This was an open-label, single-arm substudy of phase 3 COMET-TAIL (NCT04913675) assessing the safety and tolerability of a 2000 mg IV dose of sotrovimab. Symptomatic patients (aged ≥18 years) with COVID-19 at high risk for progression were enrolled from June 30 through July 11, 2022, when Omicron BA.5, BA.2.12.1, and BA.4 were the predominant circulating variants in the United States. The primary endpoint was occurrence of adverse events (AEs), serious AEs (SAEs), AEs of special interest, and COVID-19 disease-related events (DREs) through Day 8. Safety, pharmacokinetics, viral load, and hospitalization >24 hours for acute management of illness or death through Day 29 were assessed.

**Results:** All participants (n=81) were Hispanic, 58% were female, and 51% were aged ≥55 years. Through Day 8, no AEs, including infusion-related reactions or hypersensitivity, were reported; 2 participants reported DREs (mild cough, n=2). One SAE (acute myocardial infarction), which was considered unrelated to sotrovimab or COVID-19 by the investigator, occurred on Day 27 and was the only hospitalization reported. Maximum serum concentration (geometric mean) was 745.9 µg/mL. Viral load decreased from baseline through Day 29; only 2 participants (3%) had persistently high viral load (≥4.1 log_10_ copies/mL) at Day 8.

**Conclusions:** 2000 mg IV sotrovimab was well tolerated, with no new unanticipated safety signals observed.

**Key points summary:** In participants with mild to moderate coronavirus disease 2019 at risk for progression to severe disease, a 2000 mg intravenous dose of sotrovimab had a low frequency of adverse events, with no hypersensitivity, infusion-related reactions, or deaths observed.

## INTRODUCTION

Coronavirus disease 2019 (COVID-19), caused by severe acute respiratory syndrome coronavirus 2 (SARS-CoV-2), remains a serious threat to public health, with continued strain on hospitals and excess deaths due to the disease [1, 2]. Older persons and those individuals with underlying health conditions are at heightened risk of severe illness and death [3-6]. Reducing morbidity and mortality associated with COVID-19 therefore remains an important priority.

The US Food and Drug Administration (FDA) has granted Emergency Use Authorizations (EUAs) for oral antiviral therapies, including molnupiravir and nirmatrelvir/ritonavir, and has approved intravenous (IV) remdesivir for the early treatment of patients with COVID-19 in order to prevent progression to severe disease [7, 8]. The National Institutes of Health (NIH) COVID-19 guidelines recommend nirmatrelvir/ritonavir and remdesivir as the preferred first- and second-line therapies, respectively [9]. Although oral agents may be convenient with respect to administration, nirmatrelvir/ritonavir, the first line oral agent, is contraindicated for many patients due to the risk of drug-drug interactions. In such patients molnupiravir is an alternative if a preferred therapy is not possible, however treatment must be initiated within 5 days of symptom onset [10]. Intravenous remdesivir is also preferred therapy, but multiple logistical constrains may disrupt its required repeated IV administration over 3 days.

Monoclonal antibodies represent an alternative early treatment option for the prevention of progression to severe COVID-19, particularly in those individuals known to be at high risk [11]. The efficacy of one such therapy, sotrovimab, in nonhospitalized adults with mild to moderate COVID-19 at high risk of progressing to severe disease has been demonstrated in the COMET-ICE study [12]. This study was conducted from August 2020 through March 2021 in patients predominantly infected with wild-type Wuhan-Hu-1 virus [13]. In 1057 nonhospitalized participants with mild to moderate COVID-19, treatment with IV sotrovimab at a dose of 500 mg was associated with a 79% reduction in the risk of hospitalization >24 hours for acute management of any illness or death due to any cause through Day 29 when compared with placebo. A second study, COMET-TAIL, conducted from June to August 2021, compared 500 mg IV sotrovimab with intramuscular (IM) administration of 250 mg or 500 mg sotrovimab in adults and adolescents with mild to moderate COVID-19 at high risk for progression to severe COVID-19 [14]. That enrollment period coincided with a surge of the Delta variant of SARS-CoV-2, and this was the predominant variant detected in COMET-TAIL participants [15]. Results showed that 500 mg IM administration was noninferior to 500 mg IV administration. In both large trials, sotrovimab demonstrated a favorable safety profile, with no unanticipated new safety signals detected, when given by IV or IM routes.

The biggest challenge with respect to managing the burden of COVID-19 is the continuous evolution of SARS-CoV-2 [16]. In particular, the emergence of the Omicron variants of the virus has significantly changed the landscape, and whereas previously there had been a sequential emergence of single predominant variants, we are now in the era of a pool of codominant variants [17, 18]. As of January 2023, the most recent Omicron subvariants of concern include XBB.1.5, BQ.1, and BQ.1.1 [19], and it has been shown that some monoclonal antibodies, including bebtelovimab, tixagevimab/cilgavimab, and casirivimab, lose their ability to neutralize these subvariants *in vitro* [20]. However, sotrovimab has demonstrated a range of *in vitro* neutralization activity against the Omicron XBB.1.5, BQ.1, and BQ.1.1 variants using a pseudotyped virus assay, with an 11.3-, 28.5-, and 94-fold change in half maximal inhibitory concentration (IC_50_), respectively, relative to Wuhan-Hu-1 [21]. The NIH has indicated that COVID-19 caused by XBB.1.5, BQ.1 and BQ1.1 is likely to be resistant to bebtelovimab and tixagevimab/cilgavimab [9].

Although the FDA revoked the EUA for sotrovimab in the United States in April 2022 due to a reduced *in vitro* neutralization against Omicron BA.2, the predominant variant at that time, it continues to be used in patients at high risk of severe COVID-19 in other countries. Recent observational studies conducted in Europe suggest continued clinical effectiveness of sotrovimab through the Omicron BA.2 surge. In a study of >8000 patients in England, there were no differences in the risk of hospitalization for patients with mild to moderate COVID-19 treated with sotrovimab who were infected with the Omicron BA.1 or BA.2 SARS-CoV-2 variants [22]. A separate UK-based study demonstrated greater effectiveness of sotrovimab than molnupiravir in preventing COVID-19 hospitalization and death in nonhospitalized patients at high risk for progression during the BA.1 and BA.2 variant waves [23]. In a French patient cohort, there was no difference in the percentage of hospitalizations or deaths in patients with COVID-19 caused by BA.1 or BA.2 variants following treatment with sotrovimab [24]. Similarly, in a US observational study, reduced risk of all-cause hospitalization and mortality was associated with sotrovimab versus no monoclonal antibody treatment during the Delta, BA.1, and BA.2 variant waves [25]. A systematic literature review is underway to formally appraise the totality of the observational data for sotrovimab against Omicron BA.2 [26].

Understanding the safety and tolerability of higher doses of sotrovimab compared to those studied previously will provide useful information should a higher dose be necessary to treat patients with COVID-19 caused by a SARS-CoV-2 variant with reduced susceptibility to sotrovimab. The primary objective of this substudy of COMET-TAIL was to assess the safety and tolerability of 2000 mg sotrovimab when administered IV to nonhospitalized adults with mild to moderate COVID-19 at risk for progression to severe COVID-19. Exploratory objectives included assessment of clinical outcomes and antiviral activity.

## METHODS

### Participants

Participants were eligible for inclusion in the COMET-TAIL substudy (ClinicalTrials.gov Identifier: NCT04913675) if they were aged ≥18 years at the time of consent and at high risk for progression of COVID-19 based on the presence of ≥1 predefined risk factor. For participants aged 18 to 54 years, these included diabetes requiring medication, obesity (defined as body mass index ≥30 kg/m^2^), chronic kidney disease (estimated glomerular filtration rate <60 mL/min/1.73 m^2^ by Modification of Diet in Renal Disease criteria), congenital heart disease, congestive heart failure (New York Heart Association [NYHA] class II or higher), chronic lung diseases (ie, chronic obstructive pulmonary disease, moderate to severe asthma requiring corticosteroids, interstitial lung disease, cystic fibrosis, and pulmonary hypertension), sickle cell disease, neurodevelopmental disorders, immunosuppressive disease or receiving immunosuppressive medications, or chronic liver disease. Participants aged ≥55 years were eligible to participate in the study irrespective of any comorbidities.

Participants were also required to have a positive SARS-CoV-2 test result within 7 days of randomization (by any validated diagnostic test, including reverse transcriptase–polymerase chain reaction [RT-PCR] or antigen-based testing on any specimen type), oxygen saturation ≥94% on room air, and symptoms of COVID-19 (≥1 of the following: fever, chills, cough, sore throat, malaise, headache, joint or muscle pain, change in smell or taste, vomiting, diarrhea, or shortness of breath on exertion) and to have started study treatment within ≤7 days from the onset of symptoms. Participants were not eligible for the study if they were currently hospitalized, were likely to require hospitalization in the next 24 hours, or had severe COVID-19 (respiratory distress or required hospitalization for oxygen supplementation). Receiving convalescent plasma from a patient who had recovered from COVID-19 or an anti–SARS-CoV-2 monoclonal antibody within the preceding 3 months was not permitted. Additionally, antiviral treatment with nirmatrelvir/ritonavir, remdesivir, molnupiravir, or passive antibody therapies were not allowed during the study unless given as local standard of care.

### Study Design and Treatment

Details of the primary COMET-TAIL study have been reported previously [14]. The primary objective of this open-label, nonrandomized, single-arm substudy of COMET-TAIL was to describe the safety and tolerability of high-dose (2000 mg) IV sotrovimab, with an optional arm up to 3000 mg IV. The substudy enrolled participants at 7 sites in the United States from June 30 to July 11, 2022. Given the positive efficacy data from previous studies of sotrovimab in preventing disease progression in high-risk participants [12, 14], it was considered unethical to include a placebo comparator group. The study is ongoing with participants remaining under follow-up through Week 36. An initial 32 enrolled participants comprised a safety lead-in group, who received sotrovimab per study protocol and for whom relevant safety data was reviewed. An additional 49 participants were then enrolled and treated. Enrollment was discontinued early in the context of evolving variants.

Following screening, all treated participants received open-label 2000 mg sotrovimab infused IV over 60 minutes on Day 1 with follow-up through 36 weeks. Based on preliminary clinical pharmacology modeling at the time of protocol development, a 2000 mg IV dose of sotrovimab was predicted to maintain serum concentrations ≥120 μg/mL in 90% of participants for 28 days after dosing. These concentrations are ≥111-fold above wild-type tissue-adjusted 90% maximal effective concentration (EC_90_; assuming a sotrovimab lung:serum ratio of 0.25) or ≥45-fold above tissue-adjusted EC_90_ (assuming a lung:serum ratio of 0.10). Therefore, following administration of 2000 mg IV sotrovimab, serum levels are expected to remain sufficiently above the EC_90_ to provide protection against some SARS-CoV-2 variants with reduced *in vitro* neutralization potency.

Participants in the safety lead-in group were monitored for 2 hours following sotrovimab administration, with vital signs assessments predose and at 15 minutes, 30 minutes, 45 minutes, 1 hour, and 2 hours after infusion. Participants in the remainder of the cohort were monitored at similar intervals for up to 1 hour. Through Day 29, further active monitoring was conducted on an outpatient basis, starting with weekly clinic visits, including physical examination, vital signs, laboratory assessments, and collection of nasopharyngeal swabs for virologic analyses. The study is ongoing and has additional clinic visits at Weeks 12, 20, and 24. Sparse pharmacokinetic (PK) samples were collected on Day 1 (predose and at the end of the infusion) and on Days 3, 5, 8, 15, and 29. Sotrovimab serum concentrations were determined using a validated electrochemiluminescent method with a lower limit of quantification of 0.1 µg/mL. Additional samples for PK analyses are being collected through Week 24. At Weeks 8, 16, 28, 32, and 36, participants continue to be monitored via phone calls to assess for adverse events (AEs), disease-related events (DREs), serious AEs (SAEs), and the incidence and severity of any subsequent COVID-19 illness after sotrovimab dosing.

### Patient Consent Statement

The study was conducted according to the Declaration of Helsinki, Council for International Organizations of Medical Sciences international ethics guidelines, and Good Clinical Practice. The study protocol was approved by the institutional review boards/independent ethics committees at each participating study site. All participants provided written informed consent at the screening visit.

### Study Outcomes

The primary endpoint of the substudy was the occurrence of AEs, SAEs, AEs of special interest, and DREs through Day 8. All AEs were classified according to *Medical Dictionary for Regulatory Activities* (MedDRA) terminology. Secondary endpoints included the occurrence of AEs, AEs of special interest, and DREs through Week 12 and SAEs through Week 36; the incidence and titers of serum antidrug antibody (ADA) and neutralizing antibody to sotrovimab through Week 24; and serum sotrovimab PK through Week 24. AEs of special interest included infusion-related reactions (including hypersensitivity reactions occurring ≤24 hours after infusion), hypersensitivity reactions occurring at any time following dosing, immunogenicity-related AEs, and AEs potentially related to antibody-dependent enhancement of disease. This manuscript reports safety, viral load, and PK data collected through Day 29.

Several exploratory endpoints were also included to evaluate the efficacy of 2000 mg IV sotrovimab. These were progression of COVID-19 through Day 29 as defined by hospitalization >24 hours for acute management of illness due to any cause or death; or visit to a hospital emergency room for management of illness or hospitalization for acute management of illness for any duration and for any cause or death; development of severe COVID-19 requiring respiratory support including oxygen at Days 8, 15, 22 and 29; and the incidence of participants requiring intensive care unit (ICU) stay or mechanical ventilation through Day 29. Viral load in nasal secretions was assessed using quantitative RT-PCR and evaluated as the change from baseline in viral load at Days 3, 5, 8, 11, 15, 22, and 29; undetectable SARS-CoV-2 in nasal secretions at Days 3, 5, 8, 11, 15, 22, and 29; and the proportion of participants with a persistently high SARS-CoV-2 viral load (≥4.1 log_10_ copies/mL) at Day 8.

### Sample Size and Statistical Analyses

A sample size of 200 participants was initially planned to assess the safety of 2000 mg IV sotrovimab. This sample size provided a 90% probability of observing ≥1 particular AE of interest, if the true incidence of that AE was not below 1.14%, which was the frequency of infusion-related reactions observed in the COMET-ICE study.

The safety analysis set comprised all participants who were enrolled and exposed to study treatment. All analyses were conducted on this population except virologic analyses, which were conducted on all enrolled participants with a laboratory-confirmed quantifiable nasopharyngeal swab on Day 1. All participants in the safety analysis set had ≥1 nonmissing PK assessment and were included in the PK analyses.

All statistical analyses were descriptive, and no formal statistical testing was conducted. Viral load measurements reported below the lower limits of detection or quantification were imputed to 1.78 log_10_ copies/mL. All statistical analyses were performed using SAS, version 9.4. (SAS Institute Inc., Cary, NC, USA).

## RESULTS

### Participants

Due to early discontinuation of enrollment, only 81 participants received 2000 mg IV sotrovimab from the planned sample of 200 (**Table 1**; **Figure 1**). A total of 83 participants were screened; 1 failed screening, and another was not treated due to early study discontinuation. All participants were enrolled in the United States at 7 sites, with 6 in Florida. At the time of the data cutoff (October 4, 2022), 80 participants remained in the study and 1 participant had withdrawn.

**Table 1.** Baseline Demographic and Clinical Characteristics.

| Characteristic | 2000 mg IV sotrovimab (n=81) |
| --- | --- |
| Sex, n (%) |  |
| Male | 34 (42) |
| Female | 47 (58) |
| Age, mean (SD), years | 51.6 (14.6) |
| Age category, years, n (%) |  |
| <55 | 40 (49) |
| 55–64 | 26 (32) |
| 65–74 | 12 (15) |
| 75–84 | 2 (2) |
| ≥85 | 1 (1) |
| Ethnicity, n (%) |  |
| Hispanic or Latino | 81 (100) |
| Not Hispanic or Latino | 0 |
| Race, n (%) |  |
| White | 77 (95) |
| Black/African American | 4 (5) |
| BMI, mean (SD), kg/m<sup>2</sup> | 32.1 (5.2) |
| BMI category, kg/m<sup>2</sup>, n (%) |  |
| <30 | 27 (33) |
| 30 to <35 | 24 (30) |
| ≥35 | 30 (37) |
| Conditions as a risk factor for progression to severe COVID-19, n (%) |  |
| Any condition | 81 (100) |
| Obesity | 54 (67) |
| Age ≥55 years | 41 (51) |
| Chronic lung diseases | 19 (23) |
| Diabetes | 13 (16) |
| Number of conditions met, n (%) |  |
| 1 | 43 (53) |
| 2 | 30 (37) |
| 3 | 8 (10) |
| >3 | 0 |
| Received any COVID-19 vaccination, n (%) | 50 (62) |
| Fully vaccinated | 43 (53) |
| Symptom duration, days, n (%) |  |
| ≤3 | 51 (63) |
| 4–5 | 26 (32) |
| >5 | 4 (5) |

BMI, body mass index; COVID-19, coronavirus disease 2019; IV, intravenous; SD, standard deviation

**Figure 1.**
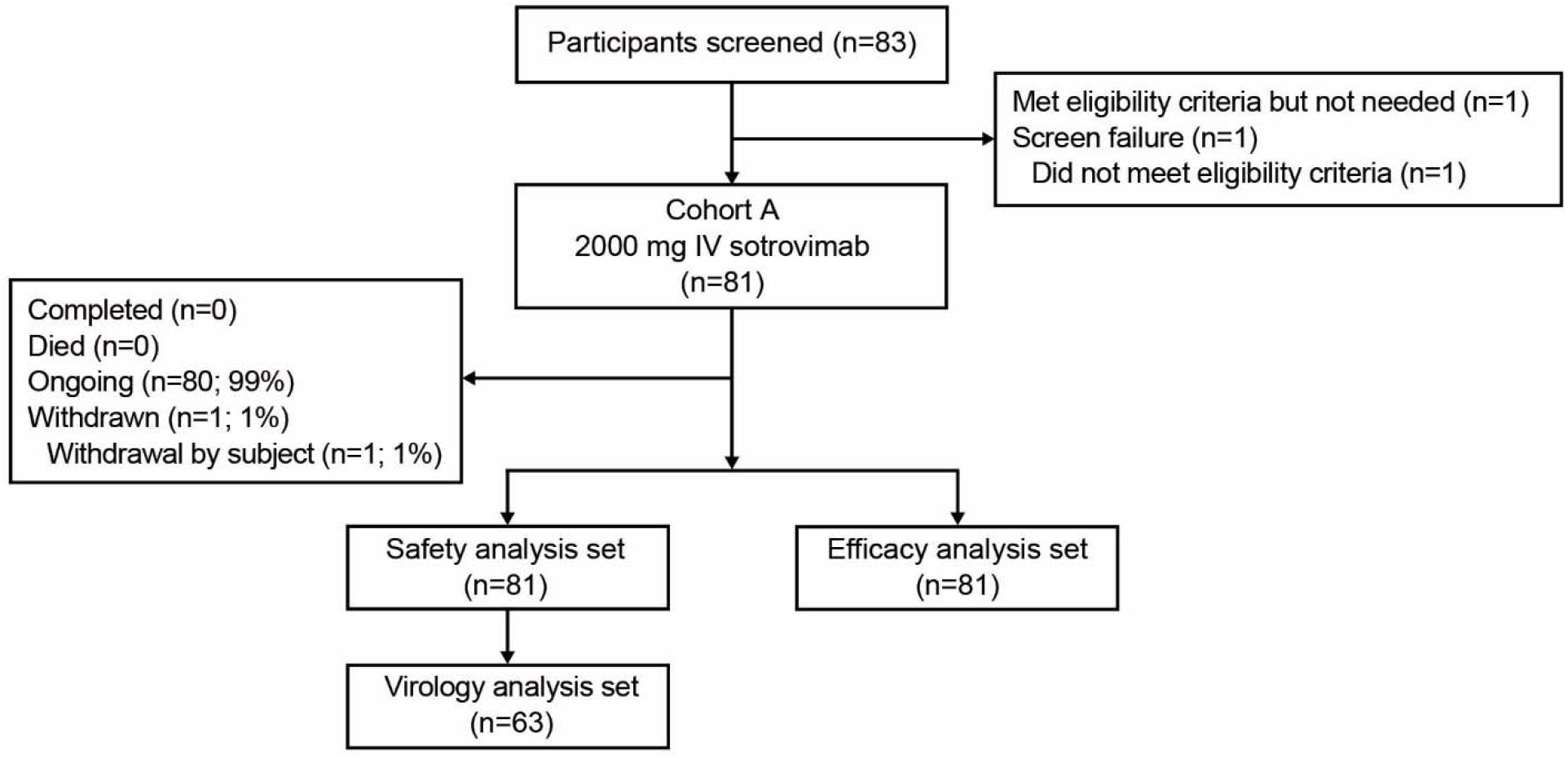
Patient enrollment and treatment assignment. IV, intravenous.

All participants were Hispanic or Latino and 77 (95%) were White; the mean age was 51.6 years, 41 (51%) participants were aged ≥55 years, and 47 (58%) participants were female. A total of 15 participants (19%) were aged ≥65 years and 38 (47%) had ≥2 risk factors for progression to severe COVID-19. None had chronic kidney disease, chronic liver disease, congenital heart disease, congestive heart failure (NYHA class II or higher), immunosuppressive disease, neurodevelopmental disorder, or sickle cell disease. All participants were immunocompetent and 50 (62%) had received ≥1 dose of a COVID-19 vaccine.

### Primary and Secondary Endpoints

For the primary endpoint, no AEs, SAEs, or AEs of special interest were observed through Day 8 (**Table 2**). A total of 2 participants experienced DREs through Day 8, including 2 cases of grade 1 cough (of 3 and 6 days’ duration, respectively). These events resolved during the study and were not associated with study discontinuation.

**Table 2.** Primary, Secondary, and Key Exploratory Endpoints.

|  | Day 8 (n=81) | Day 29 (n=81) |
| --- | --- | --- |
| Any AE, n (%) | 0 | 1 (1.2) |
| AEs related to study treatment | 0 | 0 |
| AEs leading to study treatment discontinuation,<br>dose interruption, or delay | 0 | 0 |
| Maximum grade 3/4 AEs | 0 | 1 (1.2) |
| Any SAEs | 0 | 1 (1.2) |
| SAEs related to study treatment | 0 | 0 |
| Fatal SAEs | 0 | 0 |
| DRE | 2 (2.5) | 4 (4.9) |
| Progression of COVID-19 through Day 29, n (%) | 0 | 0 |
| Hospitalization >24 hours for acute management<br>of illness due to any cause | 0 | 1 (1.2) |
| Death | 0 | 0 |
| Visit to a hospital emergency room for<br>management of illness or hospitalization for acute<br>management of illness for any duration and any<br>cause | 0 | 1 (1.2) |
| Development of severe and/or critical respiratory<br>COVID-19 as manifested by requirement for<br>respiratory support (including oxygen) | 0 | 0 |
| ICU stay or mechanical ventilation | 0 | 0 |
| Missing | 0 | 1 (1.2) |

AE, adverse event; COVID-19, coronavirus disease 2019; DRE, disease-related event; ICU, intensive care unit; SAE, serious adverse event.

For the secondary endpoint (reported through Day 29), only 1 SAE occurred, which was a grade 3 acute myocardial infarction in a participant with multiple cardiac risk factors, including old age (50 years), male sex, type 2 diabetes mellitus, hypertension, and smoking. The SAE occurred at 27 days after sotrovimab administration and was not considered related to sotrovimab or COVID-19. This SAE was reported as resolved within 3 days, and the participant continued in the study. Two participants experienced DREs through Day 29, including 2 cases of grade 3 lipase elevation and 1 case of grade 2 insomnia. The lipase elevations resolved, the insomnia was resolving at the data cutoff, and none of these events led to study discontinuation. Assessments of AEs, AEs of special interest and DREs through Week 12 and SAEs through Week 36 are ongoing.

For the secondary endpoint assessing PK after a 60-minute 2000 mg IV infusion of sotrovimab, the geometric mean (%coefficient of variation [CV]) maximum serum concentration (C_max_) was 745.9 µg/mL (41.7) and the geometric mean (%CV) area under the plasma concentration-time curve from Day 1 through Day 29 was 7696 day*µg/mL (34.6). The geometric mean (%CV) of the serum concentration at Day 29 was 203.4 µg/mL (35.5). The sotrovimab serum concentration-time plot through Day 29 is displayed in **Figure 2**.

**Figure 2.**
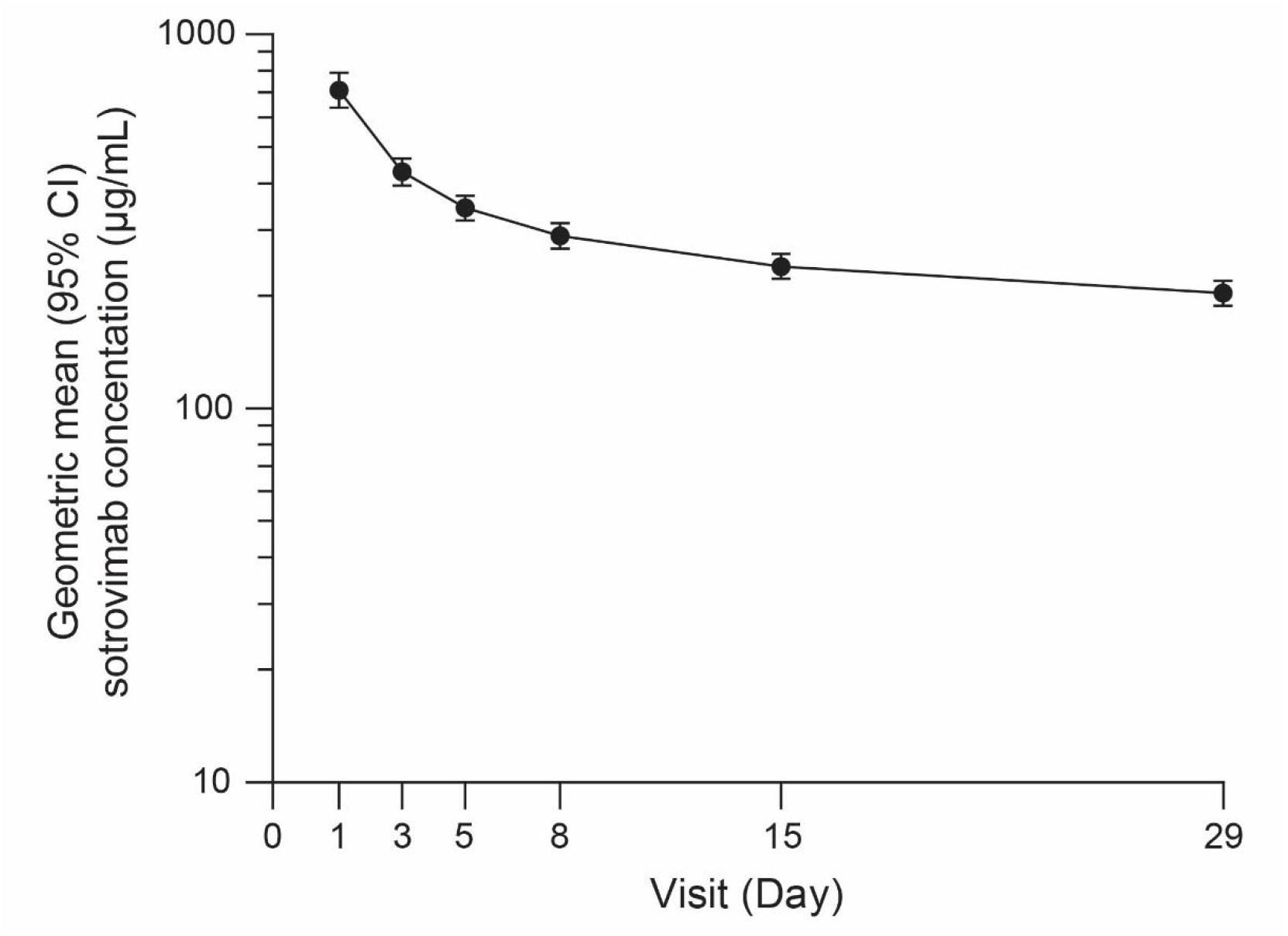
Geometric mean (95% CI) sotrovimab serum concentration-time profile after administration of 2000 mg IV sotrovimab. CI, confidence interval; IV, intravenous.

### Exploratory Endpoints

For the exploratory endpoints of progression of COVID-19 through Day 29 as assessed by hospitalization >24 hours for acute management of illness due to any cause or death; or visit to a hospital emergency room for management of illness or hospitalization for acute management of illness for any duration and for any cause or death, only the participant with the unrelated SAE of acute myocardial infarction required hospitalization. No participants experienced progression to severe and/or critical respiratory COVID-19 or ICU stay or mechanical ventilation through Day 29.

Among participants in the virology population (n=63), the mean (standard deviation) viral load at baseline was 5.02 (1.44) log_10_ copies/mL, had decreased to 2.25 (0.75) log_10_ copies/mL at Day 8, and continued to decline through Day 29 (**Figure 3A**). The proportion of participants with an undetectable SARS-CoV-2 viral load was 35% at Day 5 and consistently increased over time, rising to 45% of participants at Day 8, 81% of participants at Day 15, and 94% of participants at Day 29 (**Figure 3B**). Only 2 participants (3%) in the virology population demonstrated a persistently high SARS-CoV-2 viral load (≥4.1 log_10_ copies/mL) at Day 8.

**Figure 3.**
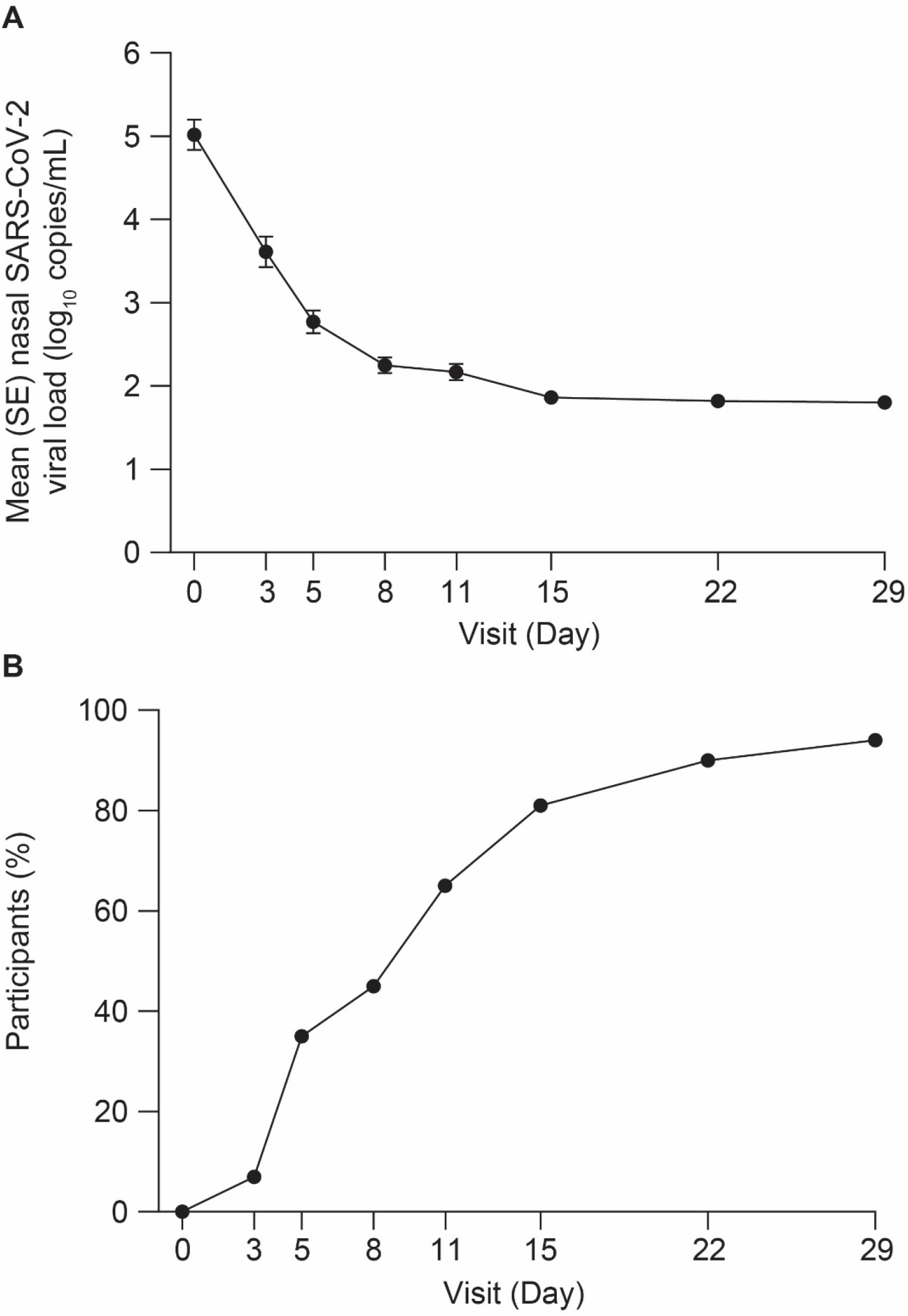
Mean (SE) nasal SARS-CoV-2 viral load (A) and percentage of participants with undetectable SARS-CoV-2 in nasal secretions (B) through Day 29. SARS-CoV-2, severe acute respiratory syndrome coronavirus 2; SE, standard error.

## DISCUSSION

This substudy of COMET-TAIL showed that a high dose (2000 mg) of sotrovimab administered IV over 60 minutes to participants with mild to moderate COVID-19 was well tolerated, with no new safety signals through Day 29 relative to the studies using the 500 mg dose.

No AEs or AEs of special interest were reported through Day 8; only 1 SAE occurred at Day 27 after dosing, which was deemed by the investigator to be unrelated to sotrovimab or COVID-19. Safety data through Weeks 12 and 36 (secondary endpoint) will be reported in the future.

There were no cases of progression, including COVID-19 hospitalization or death or development of severe COVID-19, reported during the study. Clearance of SARS-CoV-2 was observed in almost all participants by Day 29 following sotrovimab administration, with most of the viral load decline (mean, 2.78) seen by Day 8. This is of particular interest as enrollment was conducted during a period where the Omicron subvariants BA.2.12.1, BA.4, and BA.5 were the predominant circulating variants of SARS-CoV-2 (June 30 through July 11, 2022). Although *in vitro* studies suggest a moderate reduction in the neutralization of Omicron BA.2 by sotrovimab (16-fold change in IC_50_ relative to Wuhan-Hu-1) [27, 28], recent observational studies in Europe suggest maintained clinical effectiveness in cases of COVID-19 caused by this variant [22-24]. Additionally, an observational study conducted in the United States reported no decrease in effectiveness during BA.1 and BA.2 waves relative to the Delta wave [25]. Compared to Omicron BA.2, sotrovimab demonstrates similar *in vitro* neutralization potency against the Omicron subvariants BA.2.12.1, BA.4, and BA.5 (16.6-, 21.3-, and 22.6-fold change in IC_50_, respectively, relative to Wuhan-Hu-1 using a pseudotyped virus assay) [27]. Nevertheless, sotrovimab seemed to maintain clinical effectiveness against the most probable variants causing COVID-19 in the study participants.

Moving forward, it is anticipated that other emerging variants [19] and treatments providing adequate activity against these and future variants will be increasingly important. Sotrovimab has demonstrated neutralization activity *in vitro* against the recent Omicron XBB.1.5, BQ.1, and BQ.1.1 variants with a range in potency that is similar or reduced relative to BA.2 [21]. This clinical experience with sotrovimab 2000 mg and the virology data reported, together with real-world evidence data, suggest that at higher doses, sotrovimab may be useful in treating patients infected with variants for which sotrovimab has reduced neutralization activity relative to Wuhan-Hu-1. Additionally, the preliminary PK results from our study are consistent with the expectations for a 2000 mg IV dose, under assumptions of dose proportionality.

There are a number of limitations to the current study. Specifically, a single-arm without a placebo control design with a small sample size, and almost all the participants were enrolled at sites in Florida (United States) over a period of 2 weeks, leading to less heterogeneity in the SARS-CoV-2 variants than those that may have been observed with a broader geographic sample. Furthermore, there was limited racial and ethnic diversity among the study population, and no immunocompromised participants were included. The early closure of enrollment and the resultant small sample size decreased the probability of observing an AE of interest with a true incidence of 1.14% from 90% to 60%. Although the virologic analyses provide useful information with respect to the antiviral effects of high-dose sotrovimab, no firm conclusions regarding efficacy can be drawn as these outcomes were exploratory in nature and there was no control comparator.

Further findings from this study will be reported as the data become available, including safety through Weeks 12 and 36, incidence and titers of ADAs, identification of emergent viral mutants, and PK of sotrovimab after the 2000 mg IV dose through Week 24. A 3000 mg IV dose of sotrovimab is already being evaluated in healthy volunteers (COSMIC; ClinicalTrials.gov Identifier: NCT05280717) to support clinical doses and alternative infusion rates for potential future clinical studies [29]. Ongoing real-world studies outside of the United States will continue to inform a broader picture of sotrovimab activity against emerging SARS-CoV-2 variants.

Although the need for IV infusion may present as a challenge to treatment with monoclonal antibodies in the community setting during the COVID-19 pandemic [30, 31], some hospitals have created protocols to bring patients with COVID-19 into infusion centers [32, 33]. Infusion centers may also be positioned in various locations, including ambulatory centers, emergency departments, skilled nursing facilities, and residential facilities [34, 35]. Ultimately, however, the length of infusion dictates the throughput of patients requiring monoclonal antibody treatment for COVID-19 [33]. Thus, agents with shorter infusion times, or agents that may be administered non-IV (ie, IM or subcutaneous) are needed to increase access to care in patients with COVID-19.

In conclusion, sotrovimab, when administered at a dose of 2000 mg IV infused over 60 minutes, was safe and well tolerated, with a low frequency of AEs (none of which were considered related to sotrovimab) and no hypersensitivity, infusion-related reactions, or deaths in participants with mild to moderate COVID-19. These findings, together with those from other studies of sotrovimab, show that no unanticipated safety signals have been observed to date with sotrovimab at doses up to 2000 mg IV.

## Data Availability

The data collected for this study will not be made available to others.

## ACKNOWLEDGMENTS

The authors thank Jeanne McKeon, PhD, of Lumanity Scientific Inc., for medical writing support, which was funded by Vir Biotechnology, Inc., and GSK. The authors also thank Joseph Hogan, MS, Mimi Chae, MS, and Emma Gierman, MPH, of Vir Biotechnology, Inc., for clinical operations support.

## AUTHOR CONTRIBUTIONS

MT, MA, EA, AS, GS, AE-Z, JS, and SP contributed to study conception and design. GA, JM, EJ, JMG, RH-L, JD, RH, MT, and SP contributed to data analysis. MT, EA, AS, GS, AE-Z, JS, SS, CX, and SP made substantial contributions to analysis and interpretation of data. All authors were involved in drafting the article and/or revising it critically for important intellectual content, and all authors approved of its final version.

## DATA AVAILABILITY

The data collected for this study will not be made available to others.

